# AI-Assisted Tissue-Based Antibody Interpretation in Autoimmune Encephalitis: Prospective Validation and Reader Study

**DOI:** 10.64898/2026.10.08.26365042

**Authors:** Zijian Cao, Donglin Xie, Sijia Zou, Menghan Li, Dongrui Li, Pilar Martinez-Martinez, Shenghua Zong, Zongjiu Zhang

**Author notes:** These authors contributed equally to this work (Zijian Cao, Donglin Xie). Corresponding authors: Shenghua Zong; Zongjiu Zhang.

## Abstract

Tissue-based assays provide broad screening for neural autoantibodies but remain vulnerable to technical and reader-dependent variability. We evaluated an integrated workflow combining automated 3,3′-diaminobenzidine tissue-based assay processing, digital whole-slide imaging, and interpretable deep learning. Model development used 2,357 retrospective serum and cerebrospinal fluid images referred from more than 80 institutions and processed in a centralized laboratory, with reference labels established by expert-consensus tissue-based assay interpretation. A prospective validation cohort included 200 images from 200 unique patients (154 positive and 46 negative; 100 serum and 100 cerebrospinal fluid). The same images were evaluated by two expert and two novice readers in a fixed-order two-round study, with unaided interpretation followed by artificial-intelligence-assisted interpretation after a washout of at least two weeks. The standalone binary classifier achieved an area under the receiver operating characteristic curve of 0.922, accuracy of 86.5%, sensitivity of 85.7%, and specificity of 89.1% in prospective validation. Pooled reader accuracy was 90.4% in the unaided round and 97.4% in the artificial-intelligence-assisted round (McNemar P < 0.001), with specificity of 82.6% and 99.5%, respectively. Novice-reader accuracy was 83.3% unaided and 96.5% with artificial-intelligence assistance (P < 0.001), approaching unaided expert accuracy of 97.5%. Among tissue-based-assay-positive cases, reader accuracy for cytoplasmic/nuclear antigen-associated versus neural surface antigen-associated staining-pattern subclassification was 88.8% unaided and 94.0% with artificial-intelligence assistance (P < 0.001). These findings support further multicentre, cross-platform evaluation of artificial-intelligence-assisted neural autoantibody tissue interpretation.

**Abbreviated summary:** Cao et al. report prospective validation of an automated tissue-based neural autoantibody workflow combining digital imaging and interpretable deep learning. Artificial-intelligence-assisted reading was associated with higher interpretation accuracy, particularly among less-experienced readers, supporting further multicentre validation across laboratories and imaging platforms.

## Introduction

Autoimmune encephalitis (AE) is a severe acute or subacute neurologic syndrome with an estimated prevalence of 13.7 per 100,000 persons and an incidence comparable to infectious encephalitis.^1^ Diagnostic error and delay remain clinically important; in a multicentre series of adults misdiagnosed with AE, one in five experienced morbidity related to unnecessary immunotherapy.^2^ Neural autoantibody testing in serum and cerebrospinal fluid (CSF) is therefore central to the diagnostic evaluation.^3^ Antigen-specific cell-based assays (CBAs) provide targeted confirmation, whereas tissue-based assays (TBAs) using rodent brain sections provide broad screening in native tissue context and may reveal reactivities not represented in fixed antigen panels.□□□

TBA signals can be visualized by indirect immunofluorescence (IIF) or by chromogenic immunohistochemistry using 3,3′-diaminobenzidine (DAB). Commercial IIF-TBA platforms are available, whereas in-house brain-tissue assays remain common in specialized neuroimmunology laboratories□□□□. These formats differ in substrate preparation, staining procedures, detection chemistry, and image interpretation. In the present study, DAB-TBA refers specifically to a chromogenic immunohistochemistry-based tissue assay. DAB-TBA provides a permanent bright-field signal with preserved neuroanatomical context, but the workflow remains technically demanding: assay processing can vary across batches, and readers must distinguish specific brown precipitates from nonspecific background across multiple neuroanatomical regions.

Interpretation expertise is therefore a major determinant of TBA performance. Recent evaluations of commercial IIF-TBAs in autoimmune neurologic disorders documented false-negative, false-positive, and discordant interpretations and highlighted greater difficulty among less-experienced raters.□□□ In addition to binary positive/negative interpretation, tissue staining may be categorized broadly as cytoplasmic/nuclear antigen-associated (CNA) or neural surface antigen-associated (NSA), a morphologic distinction that can help guide subsequent review and antigen-specific testing. Together, these technical and interpretive demands create a substantial expertise requirement for routine neural autoantibody tissue testing.

Artificial intelligence has been applied to immunofluorescence TBA images,□ but prior work has largely focused on standalone image classification. For neuroimmunology laboratories, the translational question is not only whether a model can classify images, but whether automated assay processing, digital imaging, and AI decision support can be integrated into a workflow that preserves expert-defined tissue reactivity and supports, rather than replaces, human interpretation. Prospective evaluation and direct assessment of reader performance are particularly important because the intended use is an expertise-intensive laboratory task rather than autonomous patient-level diagnosis.

We therefore developed an integrated workflow combining automated DAB-TBA processing, digital whole-slide imaging, and a two-stage deep-learning framework for TBA positive/negative classification and CNA-versus-NSA staining-pattern subclassification. The study comprised retrospective model development and internal evaluation, prospective validation using 200 images from 200 unique patients processed within the standardized workflow, and a fixed-order two-round reader study using the same prospective image cohort. We evaluated standalone model discrimination, conditional CNA-versus-NSA subclassification, end-to-end three-class performance, and TBA interpretation by expert and novice readers under unaided and AI-assisted conditions.

## Materials and methods

### Study design and research questions

This study comprised retrospective AI development and internal evaluation, prospective model validation, and a prospective fixed-order two-round reader study (Figure 1). The primary diagnostic-accuracy question was whether the Stage 1 model could distinguish expert-consensus TBA-positive from TBA-negative images in the prospective cohort. The reader-study question was whether TBA interpretation performance differed between unaided and AI-assisted rounds. Reporting was guided by STARD-AI.1□ Dataset composition and evaluation units are summarized in Table 1.

**Figure 1.**
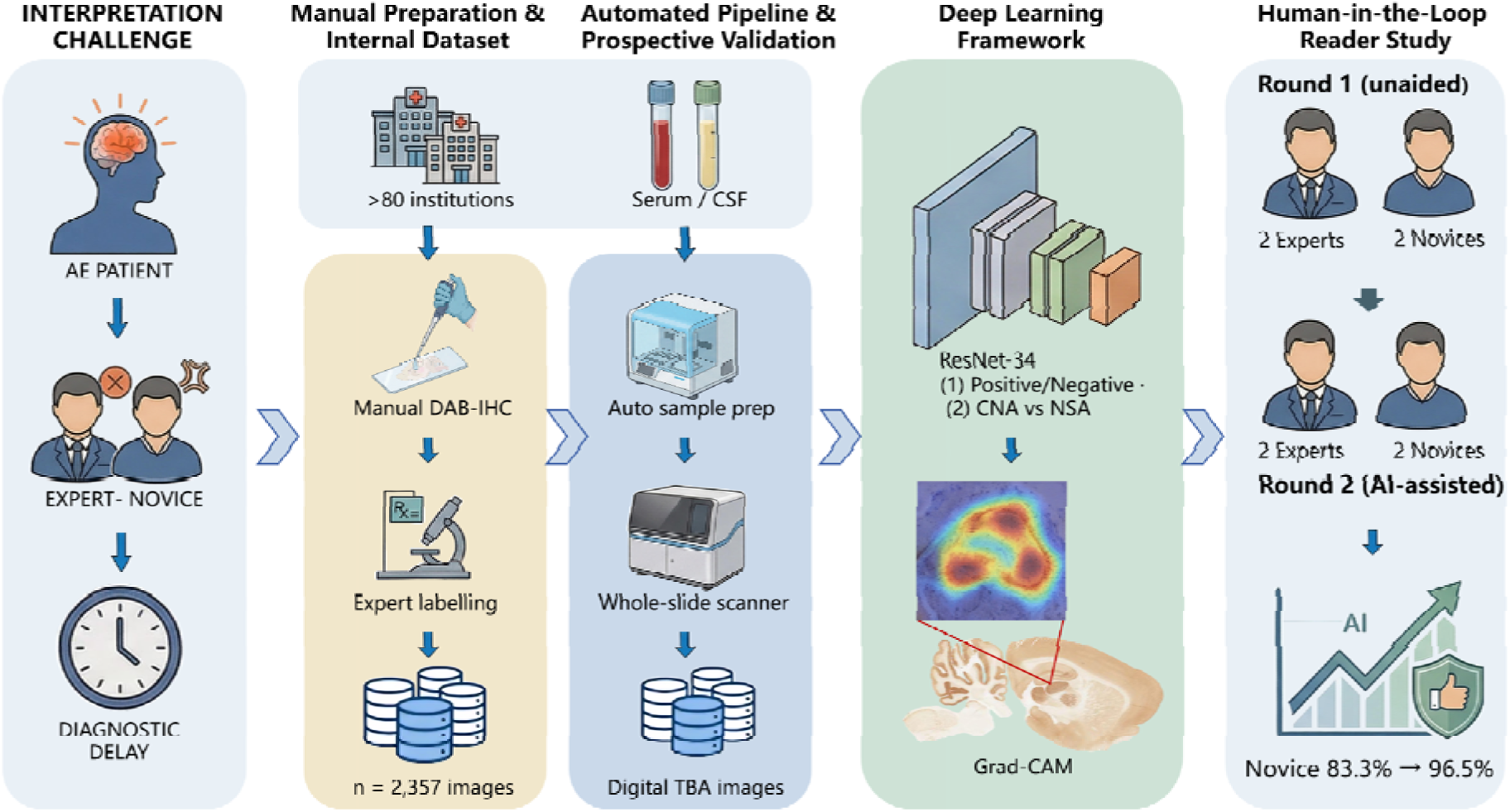
Overall study design and integrated workflow for AI-assisted DAB-TBA interpretation. The study comprised retrospective model development, prospective validation of the integrated automated workflow, and a human-in-the-loop reader study. The retrospective development dataset included 2,357 DAB-TBA images from serum and cerebrospinal fluid (CSF) specimens referred from more than 80 institutions, with reference labels established by expert-consensus TBA interpretation. Prospective specimens were processed using automated sample preparation followed by digital whole-slide imaging. A two-stage ResNet-34 framework performed TBA positive/negative classification followed by cytoplasmic/nuclear antigen-associated (CNA) versus neural surface antigen-associated (NSA) staining-pattern subclassification among TBA-positive cases, with Grad-CAM used as a qualitative interpretability aid. The prospective reader study involved two expert and two novice readers in a fixed-order, two-round design, with unaided interpretation in Round 1 followed by AI-assisted interpretation in Round 2. Novice binary TBA interpretation accuracy was 83.3% in Round 1 and 96.5% in Round 2. CNA, cytoplasmic/nuclear antigen-associated; CSF, cerebrospinal fluid; DAB-IHC, 3,3′-diaminobenzidine immunohistochemistry; Grad-CAM, gradient-weighted class activation mapping; NSA, neural surface antigen-associated; TBA, tissue-based assay.

**Table 1.** Dataset composition and evaluation structure.

| Characteristic | Training | Validation | Internal test | Prospective validation |
| --- | --- | --- | --- | --- |
| Original images, n | 1,650 | 236 | 471 | 200 |
| Total views used, n | 4,950 | 708 | 1,413 | 200 |
| Views per original image | 3 | 3 | 3 | 1 |
| Fixed image variants | Original + Gaussian blur + brightness/contrast adjustment | Original + Gaussian blur + brightness/contrast adjustment | Original + Gaussian blur + brightness/contrast adjustment | Original image only |
| TBA-positive original images, n | — | — | 132 | 154 |
| TBA-negative original images, n | — | — | 339 | 46 |
| Serum original images, n | — | — | 296 | 100 |
| CSF original images, n | — | — | 175 | 100 |
| Analysis unit for reported performance | — | 708 validation views for threshold optimisation | 1,413 inference views | 200 original images |
| Collection period | June 2024–September 2025 (retrospective) | June 2024–September 2025 (retrospective) | June 2024–September 2025 (retrospective) | October 2025 (prospective) |
| Data source | Referral-derived (>80 institutions); centralized KingMed workflow | Referral-derived (>80 institutions); centralized KingMed workflow | Referral-derived (>80 institutions); centralized KingMed workflow | Centralized laboratory workflow |
| Reference standard | Expert-consensus TBA interpretation | Expert-consensus TBA interpretation | Expert-consensus TBA interpretation | Expert-consensus TBA interpretation |

Note. The internal dataset was partitioned at the patient level before augmentation to prevent overlap between training, validation, and test subsets. Each internal original image was represented by the original image and two fixed variants generated using mild Gaussian blur and brightness/contrast adjustment. Threshold optimisation was performed using the 708 validation views. Reported internal test metrics were calculated across 1,413 inference views derived from 471 original images. The prospective validation cohort was evaluated using original images only. CBA results were complementary antigen-specific information and were not required for assignment of the TBA positive/negative reference label. CSF, cerebrospinal fluid; TBA, tissue-based assay; CBA, cell-based assay.

### Standard protocol approvals, registrations, and patient consents

Ethical approval was obtained from the Institutional Review Board of Tsinghua University (THU-01-2025-1020) and the KingMed Medical Ethics Committee (2026073). For the retrospective development dataset, written informed consent was waived because fully de-identified archival images were used. For the prospective validation cohort collected in October 2025, written informed consent was obtained from all participants or their legal representatives. The study was conducted in accordance with the Declaration of Helsinki. No identifiable participant information is presented.

### Dataset and reference standard

A total of 2,357 bright-field DAB-TBA images were retrospectively collected from the clinical archive of Nanjing KingMed Medical Laboratory between June 2024 and September 2025, representing referrals from more than 80 medical institutions across China. The archive included 1,478 serum images and 879 CSF images. Expert-consensus TBA interpretation classified 660 images as positive and 1,697 as negative. Each image was independently reviewed by at least 2 laboratory professionals with a minimum of 3 years of TBA interpretation experience; discordant interpretations were adjudicated by a senior expert. Reference-standard reviewers were blinded to all AI model outputs during label assignment and adjudication. No indeterminate reference-standard labels remained after adjudication.

Antigen-specific CBA testing using the Newterrain Autoimmune Encephalitis Autoantibody Panel IgG Detection Kit (Newterrain Biotech Co., Ltd., Tianjin, China), covering 14 neural antigens (NMDAR, LGI1, GABABR, AMPA1, AMPA2, CASPR2, IgLON5, DPPX, DRD2, mGluR1, mGluR5, GAD65, GlyR1, and Neurexin-3α), provided complementary antigen-specific information and was not required for assignment of the TBA positive/negative reference label. The internal dataset was partitioned at the patient-ID level into 1,650 training, 236 validation, and 471 internal-test original images (7:1:2), ensuring that images from the same patient did not cross subsets.

A prospective validation cohort of 200 images from 200 unique patients was collected in October 2025 and processed within the integrated automated workflow. The cohort comprised 154 expert-consensus TBA-positive and 46 TBA-negative images and was balanced by specimen type, with 100 serum and 100 CSF specimens. One original image per patient was used for prospective evaluation without augmentation.

### Automated DAB-TBA processing and digital imaging

Slides were processed using the IF-100 automated sample dispensing system (Beijing Hejie Innovation Biomedical Technology Co., Ltd., Beijing, China; Class I medical device filing no. 20250397) operated in DAB-IHC mode. Programmed steps included reagent dispensing and controlled DAB incubation. After staining, slides were digitized using a KF-PRO-400-H1 whole-slide scanner (Jiangfeng Digital Imaging Technology, China) under bright-field microscopy at ×200 effective magnification (0.5 μm/pixel) with standardized scanning parameters. The study did not include a direct manual-versus-automated processing comparison.

### Deep-learning framework and evaluation

A ResNet-34 convolutional neural network11 pretrained on ImageNet12 was used for Stage 1 TBA positive/negative classification. A second ResNet-34 model was used for Stage 2 CNA-versus-NSA staining-pattern subclassification among reference-standard TBA-positive images. In both stages, the original fully connected layer was replaced by dropout (rate 0.5) followed by a task-specific linear output layer. Input images were resized to 224 × 224 pixels and normalized using ImageNet channel-wise statistics. Standalone model predictions were generated from image data only, without access to reference-standard labels.

For the internal dataset, each original image was represented by 3 fixed views: the original image, a mild Gaussian-blur variant, and a mild brightness/contrast-adjusted variant (Supplementary Fig. 1). Thus, 1,650 training originals generated 4,950 views, 236 validation originals generated 708 views, and 471 internal-test originals generated 1,413 inference views. Patient-level partitioning was completed before generation of augmented views. The internal preprocessing pipeline additionally included random rotation (±15°), horizontal flipping (probability 0.5), and color jitter. Prospective validation used original images only.

Stage 1 used binary cross-entropy with logits loss and a positive-class weight based on the negative-to-positive training ratio. Training used AdamW (learning rate 1 × 10□□; weight decay 1 × 10□□), with early stopping after 6 epochs without validation-AUC improvement. The Stage 1 decision threshold was selected using the 708 validation views to maximize F1 and was fixed before internal-test and prospective evaluation. Stage 2 used focal loss with inverse-frequency class weighting, a 3-epoch frozen-backbone warm-up, cosine annealing, gradient-norm clipping, and early stopping based on validation macro F1. A dual-threshold grid search over 0.05-0.95 selected the Stage 2 thresholds on validation data, which were then fixed.

Gradient-weighted Class Activation Mapping (Grad-CAM)13 was integrated as a qualitative interpretability aid. During AI-assisted reading, readers were shown the categorical model prediction, a continuous model output score, and the corresponding Grad-CAM map. Grad-CAM was not treated as a validated causal explanation.

### Reader study

The same 200 prospective images were used in a fixed-order, two-round reader study. Four readers participated: 2 experts (each with ≥5 years of dedicated TBA interpretation experience) and 2 novices (each with <1 year of TBA experience). In Round 1, readers independently interpreted all images without AI assistance. After a washout interval of at least 2 weeks, the same readers completed Round 2 with AI assistance. Image order was randomized independently in each round, but reading-condition order was fixed, with unaided interpretation always preceding AI-assisted interpretation. Readers were free to weight the AI information as they judged appropriate.

### Statistical analysis

No formal sample-size calculation was performed. The analyzed datasets comprised the available retrospective archive and the prospective cohort of 200 participants. No analyzed observations had missing reference-standard labels or model outputs, and no imputation was performed. Standalone model performance was summarized using AUC, accuracy, sensitivity, specificity, positive predictive value (PPV), and F1 score, as appropriate. Internal-test metrics were calculated at the inference-view level, whereas prospective validation metrics were calculated at the original-image level. The primary reader-study comparison was the difference in overall TBA interpretation accuracy between Round 1 and Round 2, assessed using the paired McNemar test. Secondary reader outcomes included sensitivity, specificity, and Cohen κ, with κ interpreted as chance-corrected agreement with the reference labels rather than inter-reader agreement. A Bonferroni-adjusted threshold of α = 0.0125 was applied to the 4 reader-experience comparisons across the binary and CNA-versus-NSA tasks. Specimen-type subgroup analyses were exploratory; nominal 2-sided p values are reported without additional multiplicity adjustment. Analyses were performed using Python 3.10 (scikit-learn 1.3.2; SciPy 1.11.4) and R 4.3.

## Results

### Standalone model performance and prospective validation

On the retrospective internal test set, 471 original images were represented by 1,413 inference views. At the inference-view level, the Stage 1 binary classifier achieved an area under the receiver operating characteristic curve (AUC) of 0.868, accuracy of 76.9%, sensitivity of 76.5%, and specificity of 77.0% (Table 2; Figure 2A; Supplementary Fig. 2A). Dataset composition and evaluation units are summarized in Table 1.

**Figure 2.**
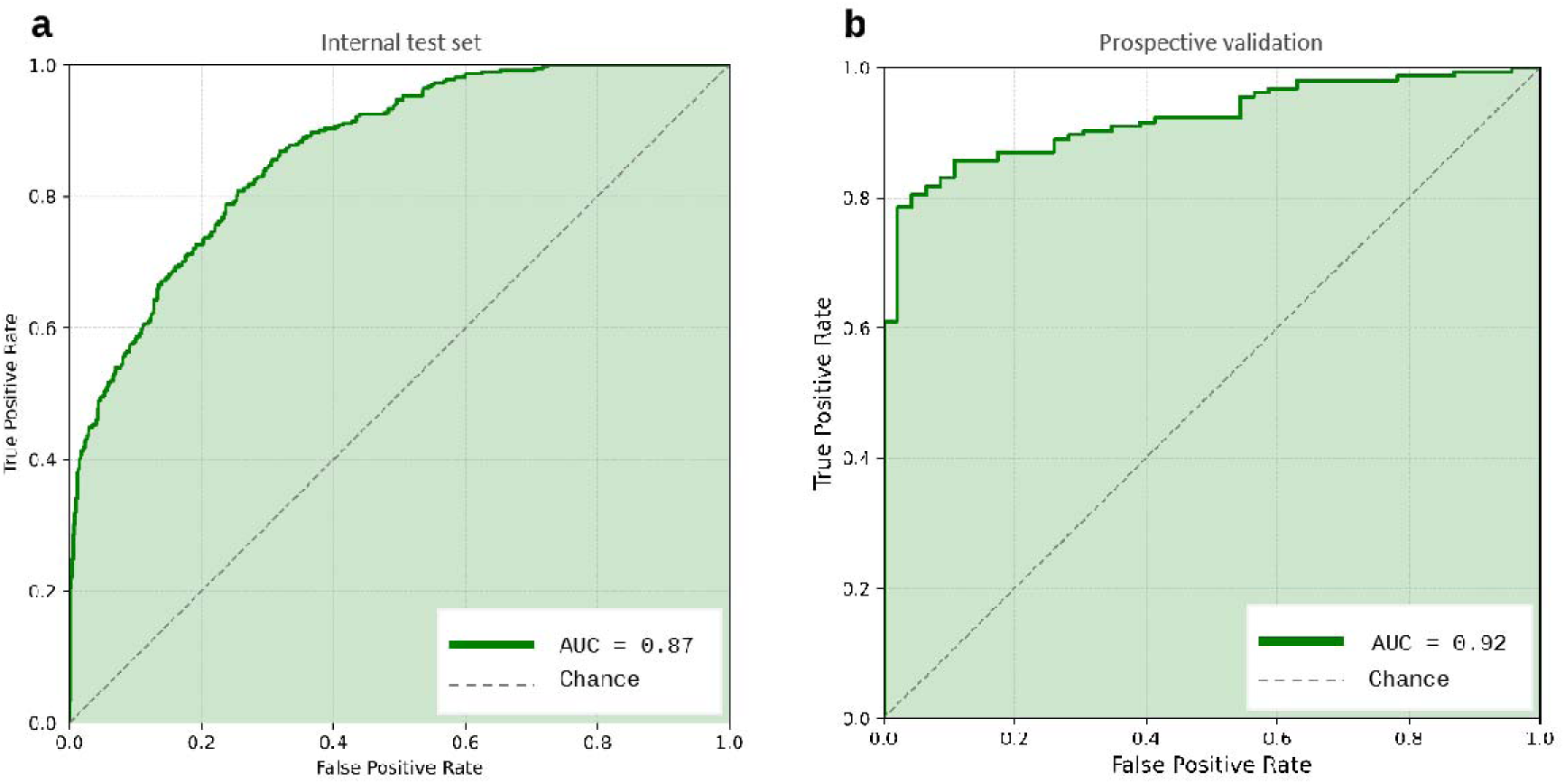
Receiver operating characteristic curves for binary TBA positive/negative classification. (A) Internal test set, comprising 1,413 inference views derived from 471 original images (AUC = 0.868). (B) Prospective validation cohort, comprising 200 original images from 200 unique patients (AUC = 0.922). The diagonal dashed line represents chance-level discrimination. Internal-test performance was evaluated at the inference-view level, whereas prospective validation was evaluated at the original-image level. AUC, area under the receiver operating characteristic curve; TBA, tissue-based assay.

**Table 2.** Binary TBA positive/negative classification performance by cohort and specimen type.

| Cohort / subgroup | n (analysis units) | Accuracy | AUC | Sensitivity | Specificity | PPV | F1 |
| --- | --- | --- | --- | --- | --- | --- | --- |
| Internal test set |  |  |  |  |  |  |  |
| Overall | 1,413 views (471 originals) | 76.9% | 0.868 | 76.5% | 77.0% | 57.0% | 0.653 |
| Serum | 888 views (296 originals) | 72.9% | 0.864 | 84.7% | 67.3% | 55.1% | 0.668 |
| CSF | 525 views (175 originals) | 87.4% | 0.949 | 76.2% | 91.8% | 78.3% | 0.772 |
| Prospective validation cohort |  |  |  |  |  |  |  |
| Overall | 200 images | 86.5% | 0.922 | 85.7% | 89.1% | 96.4% | 0.907 |
| Serum | 100 images | 92.0% | — | 96.1% | 79.2% | 93.6% | 0.948 |
| CSF | 100 images | 81.0% | — | 75.6% | 100.0% | 100.0% | 0.861 |

Note. Internal test metrics were calculated at the inference-view level, whereas prospective validation metrics were calculated at the original-image level; the two sets of estimates should therefore not be interpreted as directly equivalent. AUC was not separately calculated for prospective serum and CSF subgroups. PPV, positive predictive value; CSF, cerebrospinal fluid; TBA, tissue-based assay.

In the prospective validation cohort of 200 original images, the Stage 1 classifier achieved an AUC of 0.922, accuracy of 86.5%, sensitivity of 85.7%, specificity of 89.1%, positive predictive value of 96.4%, and F1 score of 0.907 (Table 2; Figure 2B). The prospective cohort was enriched for TBA-positive cases (154/200, 77.0%) compared with the 28.0% positive proportion in the retrospective archive. Because the internal and prospective evaluations differed in analysis unit, case mix, class prevalence, and acquisition conditions, their AUCs should not be interpreted as directly comparable estimates of cohort performance.

For end-to-end three-class inference in the prospective cohort, images classified as positive by Stage 1 were passed to Stage 2, whereas Stage 1-negative images retained the negative label. Overall accuracy across the negative, CNA, and NSA categories was 81.5% (Supplementary Table 1; Supplementary Fig. 4). All prospective slides were processed using the IF-100 system and digitized on the KF-PRO-400-H1 scanner under the standardized study workflow. Because the study did not include a direct manual-versus-automated processing comparison, the independent contribution of automated processing to model performance could not be isolated.

### Performance by specimen type and staining pattern

Within the internal test set, the model showed higher observed discrimination and specificity in CSF than in serum, but lower sensitivity. Among 525 CSF inference views derived from 175 original images, AUC was 0.949, accuracy 87.4%, sensitivity 76.2%, and specificity 91.8%. Among 888 serum inference views derived from 296 original images, AUC was 0.864, accuracy 72.9%, sensitivity 84.7%, and specificity 67.3% (Table 2; Figure 3; Supplementary Fig. 2B-C). No direct statistical comparison of serum and CSF AUCs was performed.

**Figure 3.**
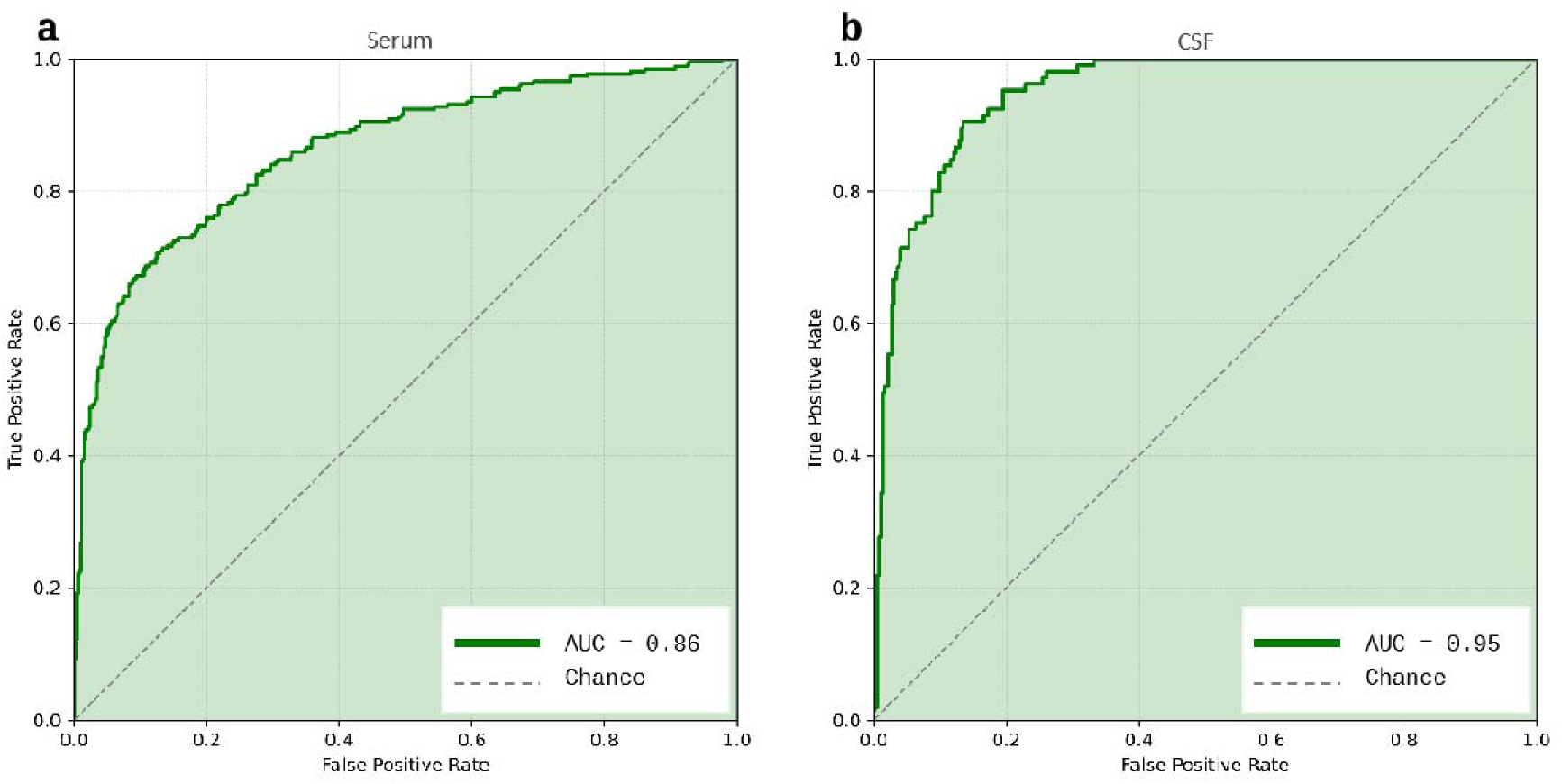
Receiver operating characteristic curves for binary TBA positive/negative classification stratified by specimen type in the internal test set. (A) Serum specimens, comprising 888 inference views derived from 296 original images (AUC = 0.864). (B) Cerebrospinal fluid (CSF) specimens, comprising 525 inference views derived from 175 original images (AUC = 0.949). The diagonal dashed line represents chance-level discrimination. The higher observed AUC in CSF indicates greater discrimination in this subgroup; no direct statistical comparison between specimen types was performed. AUC, area under the receiver operating characteristic curve; CSF, cerebrospinal fluid; TBA, tissue-based assay.

The lower serum specificity is compatible with the greater nonspecific background that can occur in serum DAB-TBA preparations. However, the study did not include paired manual-processing controls or a mechanistic assessment of specimen-type effects; the serum-CSF differences should therefore be interpreted descriptively rather than as evidence of a causal processing effect.

Among 132 reference-standard TBA-positive internal-test original images represented by 396 inference views, the conditional Stage 2 model achieved 91.7% accuracy and an AUC of 0.797 for CNA-versus-NSA staining-pattern subclassification (Supplementary Table 2; Supplementary Fig. 3). CNA sensitivity was 42.9%, whereas NSA recall was 97.5%. The lower CNA sensitivity identifies a weakness for morphologically heterogeneous or relatively scarce CNA patterns. Stage 2 should therefore be interpreted as staining-pattern subclassification rather than antigen identification; its outputs are intended to support downstream review and antigen-specific testing rather than establish pathogenicity or treatment decisions.

### AI-assisted reader study

The prospective fixed-order two-round reader study used the same 200 images evaluated in prospective model validation. Four readers completed Round 1 without AI assistance and, after a washout of at least 2 weeks, Round 2 with the categorical prediction, continuous model output score, and Grad-CAM map available as a combined decision-support interface. Image order was randomized independently in each round, whereas reading-condition order was fixed.

Overall TBA interpretation accuracy was 90.4% in Round 1 and 97.4% in Round 2 (McNemar P < 0.001). Sensitivity was 92.7% and 96.8%, specificity was 82.6% and 99.5%, and Cohen’s κ was 0.735 and 0.928 in Rounds 1 and 2, respectively (Figure 4A; Supplementary Table 3). In exploratory specimen-type analyses, accuracy was 90.0% and 95.8% for serum and 90.8% and 99.0% for CSF in Rounds 1 and 2, respectively (Supplementary Table 3).

**Figure 4.**
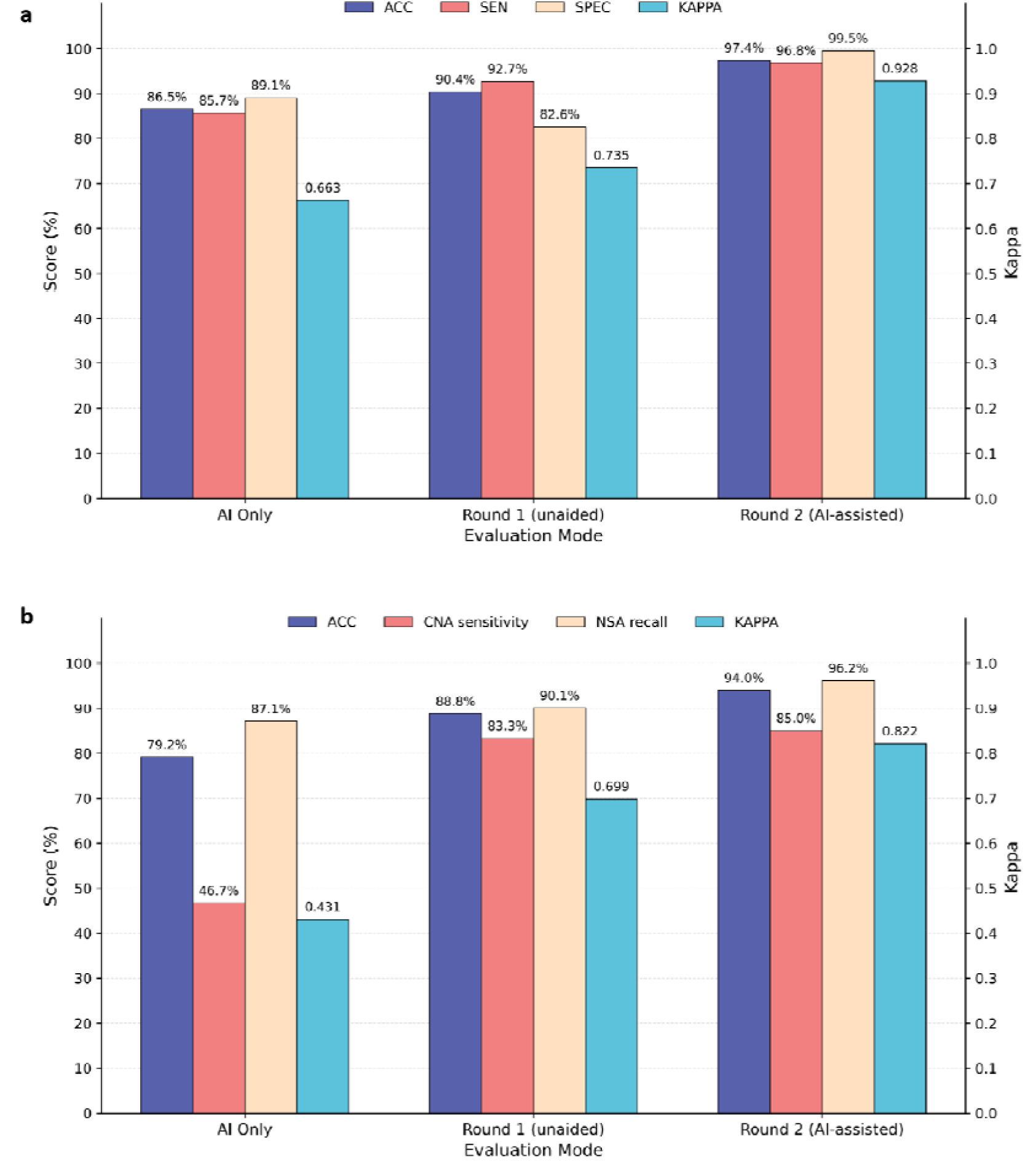
TBA interpretation performance across AI-only, unaided, and AI-assisted reading modes in the prospective reader-study cohort. (A) Binary positive/negative TBA interpretation in the overall cohort (n = 200). AI-only accuracy was 86.5%; pooled reader accuracy was 90.4% in Round 1 (unaided) and 97.4% in Round 2 (AI-assisted). Sensitivity was 92.7% and 96.8%, and specificity was 82.6% and 99.5%, respectively. (B) CNA-versus-NSA staining-pattern subclassification among TBA-positive cases (n = 154). AI-only accuracy was 79.2%; pooled reader accuracy was 88.8% in Round 1 and 94.0% in Round 2, with CNA sensitivity of 83.3% and 85.0% and NSA recall of 90.1% and 96.2%, respectively. Cohen’s κ is shown on the secondary axis. CNA, cytoplasmic/nuclear antigen-associated; NSA, neural surface antigen-associated; TBA, tissue-based assay.

The largest observed difference occurred among novice readers. Novice binary interpretation accuracy was 83.3% without AI assistance and 96.5% with AI assistance (P < 0.001), approaching the unaided expert accuracy of 97.5%. Expert accuracy was 97.5% in Round 1 and 98.3% in Round 2 (P = 0.508) (Figure 5A; Supplementary Table 5). Because only two experts and two novices participated, these experience-stratified findings should be interpreted as results from the participating readers rather than population-level estimates.

**Figure 5.**
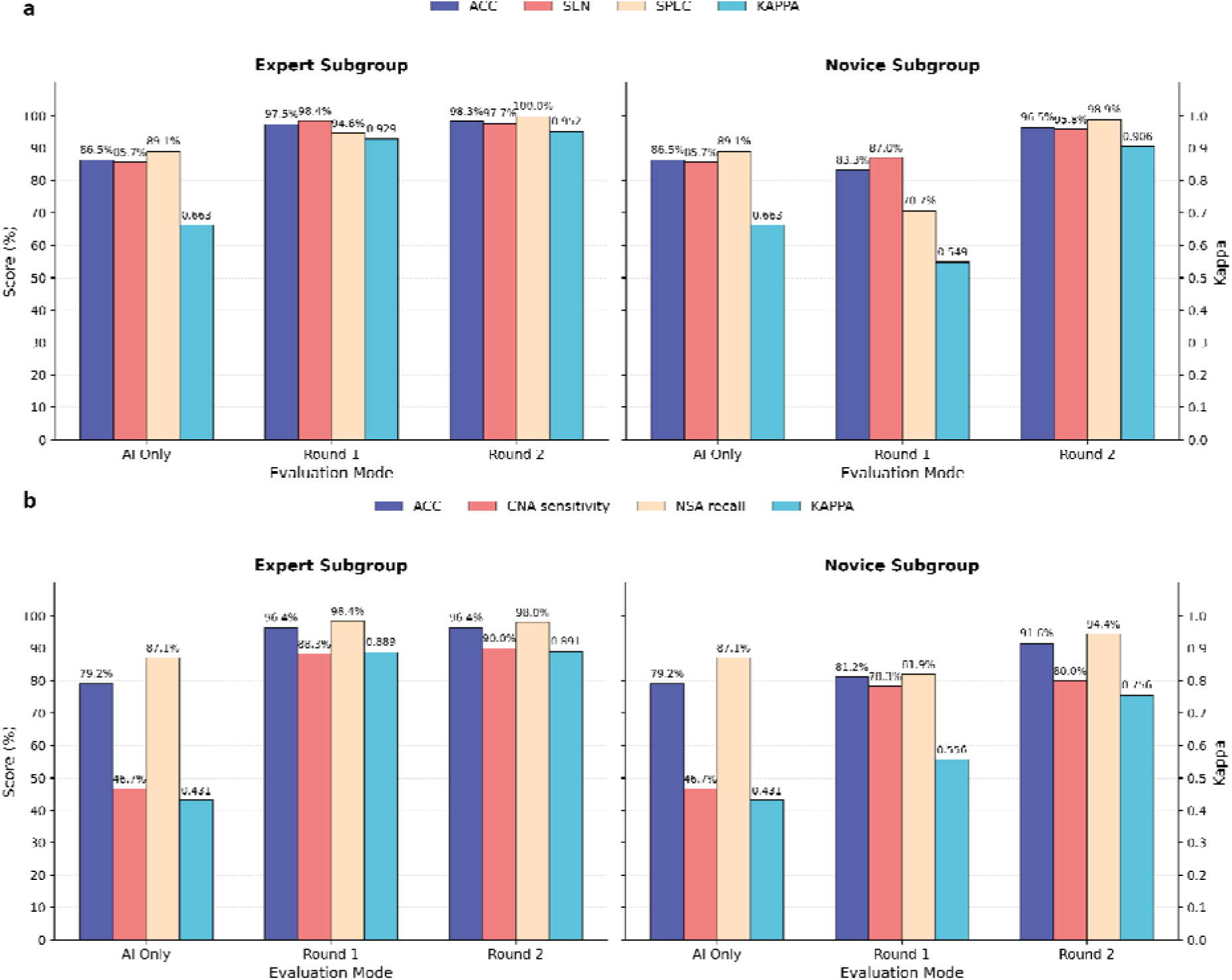
AI-assisted TBA interpretation performance according to reader experience. The upper panels show binary positive/negative TBA interpretation, and the lower panels show CNA-versus-NSA staining-pattern subclassification among TBA-positive cases. For the binary task, expert-reader accuracy was 97.5% in Round 1 and 98.3% in Round 2, whereas novice-reader accuracy was 83.3% and 96.5%, respectively. For CNA-versus-NSA subclassification, expert-reader accuracy was 96.4% in both rounds, whereas novice-reader accuracy was 81.2% in Round 1 and 91.6% in Round 2. AI-only performance is shown in each panel as a common reference and does not represent an experience-specific model. ACC, accuracy; CNA, cytoplasmic/nuclear antigen-associated; NSA, neural surface antigen-associated; SEN, sensitivity; SPEC, specificity; TBA, tissue-based assay; κ, Cohen’s kappa.

Among the 154 reference-standard TBA-positive prospective cases, reader accuracy for CNA-versus-NSA subclassification was 88.8% in Round 1 and 94.0% in Round 2 (P < 0.001); CNA sensitivity was 83.3% and 85.0%, and NSA recall was 90.1% and 96.2%, respectively (Figure 4B; Supplementary Table 4). Expert accuracy was 96.4% in both rounds (P = 1.000), whereas novice accuracy was 81.2% in Round 1 and 91.6% in Round 2 (P < 0.001) (Figure 5B; Supplementary Table 6). Exploratory specimen-type analyses showed accuracies of 89.1% and 93.4% among serum-positive cases and 88.5% and 94.6% among CSF-positive cases in Rounds 1 and 2, respectively.

### Grad-CAM interpretability

Representative Grad-CAM maps were examined qualitatively to characterize the spatial distribution of model attention. Across selected examples, model attention varied in location and extent across brain tissue sections, with some cases showing more localized activation and others showing broader or less tissue-focused patterns (Figure 6). These observations are descriptive and do not establish causal biological reasoning or independent diagnostic validity.

**Figure 6.**
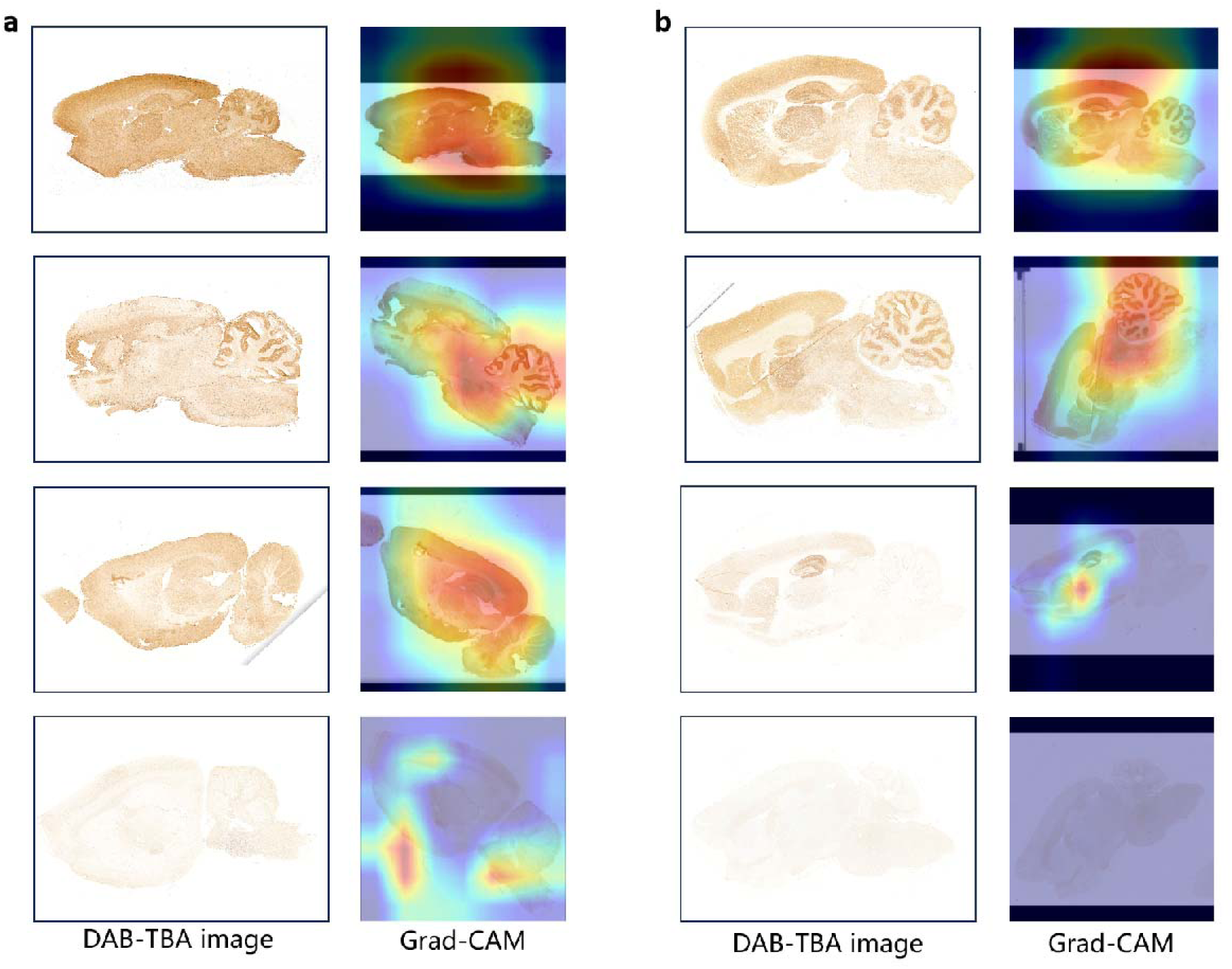
Representative spatial attention patterns visualized by Grad-CAM during DAB-TBA classification. Representative DAB-TBA images from two independent example sets are shown alongside their corresponding Grad-CAM saliency overlays. Panels a and b each contain four representative specimens illustrating heterogeneous spatial attention patterns across brain tissue sections. Warmer colors indicate image regions contributing more strongly to the model output, whereas cooler colors indicate lower relative contribution. These visualizations are presented as qualitative interpretability aids and should not be interpreted as direct evidence of causal biological reasoning or independent diagnostic validity. DAB-TBA, 3,3′-diaminobenzidine tissue-based assay; Grad-CAM, gradient-weighted class activation mapping.

## Discussion

This study developed and prospectively evaluated an integrated workflow combining automated DAB-TBA processing, digital whole-slide imaging, deep-learning classification, and reader-facing decision support for neural autoantibody tissue interpretation. The evaluation linked retrospective model development with prospective validation and a human-in-the-loop reader study, allowing the model to be assessed both as a standalone classifier and as a component of laboratory interpretation.

Three findings warrant emphasis. First, the Stage 1 model showed high discrimination in prospective validation (AUC 0.922), whereas internal-test performance was reported at the inference-view level (AUC 0.868), precluding a simple case-level comparison between cohorts. Second, reader accuracy and particularly specificity were higher in the AI-assisted round, with the largest observed difference among novice readers. Third, CNA-versus-NSA subclassification remained more difficult for CNA patterns, with low standalone CNA sensitivity despite high NSA recall. These findings suggest that the main near-term value of the framework lies in supporting binary TBA interpretation and reader decision-making rather than autonomous pattern-level classification.

The results extend recent work on TBA performance in autoimmune neurologic disorders. Two recent evaluations of commercial IIF-TBAs documented clinically relevant false-negative, false-positive, and discordant interpretations and highlighted the importance of reader experience.□□□ Our study addresses a complementary question: whether an in-house DAB-TBA workflow can be standardized digitally and paired with AI decision support. The goal is not to replace antigen-specific confirmatory testing or specialist review, but to support a broad tissue-screening step that remains dependent on morphologic expertise.

The most directly comparable AI study applied a multichannel deep-learning framework to IIF-TBA images for AE classification and reported 88.5% accuracy on an internal four-class task. Direct numerical comparison with our prospective binary-classification results is inappropriate because the imaging modality, task definition, class distribution, and validation design differ. Our study extends this line of work by evaluating DAB-TBA, incorporating prospective validation, and directly examining human interpretation with and without AI assistance. Referral material originated from more than 80 institutions, but staining, scanning, and model evaluation were centrally performed; these data therefore demonstrate broad referral provenance rather than true site-level multicentre workflow validation.

Specimen type influenced model operating characteristics. CSF showed higher observed AUC and specificity than serum in the internal test set (AUC 0.949 vs 0.864; specificity 91.8% vs 67.3%) but lower sensitivity (76.2% vs 84.7%). These differences indicate different operating characteristics rather than uniformly superior performance in CSF and should be interpreted descriptively because the study did not include a direct statistical comparison of AUCs or a mechanistic assessment of the source of the difference. The relatively lower serum specificity is compatible with the more complex nonspecific background encountered in serum TBA interpretation, but the present study was not designed to establish a causal explanation.

The low standalone CNA sensitivity (42.9%) is a more direct limitation of the current model. CNA staining encompasses morphologically heterogeneous patterns and was less well represented than NSA in the available data. The high NSA recall but limited CNA sensitivity indicates that the conditional Stage 2 model should not be used as an autonomous antigen-level classifier. Its output is better viewed as a morphologic decision-support signal that may help organize downstream review and antigen-specific testing. Expansion of morphologically diverse CNA examples will be necessary before this component can be relied upon independently.

The reader-study findings are particularly relevant to implementation because TBA interpretation is expertise-intensive. AI-assisted overall accuracy was 97.4%, compared with 90.4% during unaided reading, and specificity was 99.5% compared with 82.6%. The largest observed change was therefore a reduction in false-positive classifications. Because Round 2 combined categorical model predictions, continuous output scores, and Grad-CAM maps, the between-round difference should be attributed to the AI-assisted condition as a whole rather than to any single interface component. The study was not designed to determine whether the categorical prediction, model score, or saliency map contributed most strongly to reader decisions.

Experience-stratified analyses showed a large observed difference among the two novice readers (83.3% to 96.5%, P < 0.001), whereas the two experts showed little change (97.5% to 98.3%, P = 0.508). AI-assisted novice performance therefore approached, but did not exceed, the unaided expert performance observed in this study. This pattern suggests that decision support may be most useful when baseline TBA interpretation experience is limited. However, the comparison involved only four readers from a single laboratory and should not be generalized to all expert and novice readers without larger, multisite reader studies.

The fixed-order design also constrains causal interpretation. Unaided reading always preceded AI-assisted reading, and although image order was independently randomized and a washout interval was used, residual familiarity, learning, or period effects cannot be excluded. The observed between-round improvement therefore cannot be attributed exclusively to AI assistance. The present study also did not assess whether repeated exposure to model outputs produces a durable training or pedagogical effect. Longitudinal reader studies would be required to distinguish immediate decision support from longer-term learning.

A methodological feature of the study was the integration of automated DAB-TBA processing with digital whole-slide imaging. Programmed assay-processing and standardized scanning provided consistent technical conditions for prospective evaluation. However, the study did not include a direct manual-versus-automated reproducibility comparison, so any reduction in technical variability cannot be quantified from the present data. Likewise, the model was evaluated on a single scanner platform. Independent evaluation across laboratories, staining systems, and scanner platforms remains necessary before the workflow can be considered technically generalizable.

Several additional limitations should be considered. The prospective validation cohort was enriched for TBA-positive cases (154/200, 77.0%) and therefore did not reflect the approximately 28.0% positive proportion observed in the retrospective routine-testing archive. Consequently, prevalence-dependent measures, including overall accuracy and positive predictive value, should not be directly extrapolated to an unselected clinical testing population. Internal-test metrics were calculated across inference views derived from augmented originals, whereas prospective metrics were calculated at the original-image level. Formal demographic fairness analyses were not performed. Finally, the study evaluated TBA image interpretation rather than patient-level diagnosis, treatment decisions, laboratory turnaround time, or clinical outcomes.

Future work should prioritize prospective multicentre validation in consecutive clinical cohorts, expansion of morphologically diverse CNA-pattern cases, and evaluation across different laboratory and scanner platforms. Prospective implementation studies should also assess how AI-assisted TBA interpretation performs under routine case prevalence and workflow constraints, including effects on reader performance, false-positive classifications, turnaround time, and downstream antigen-specific testing. Larger reader studies could clarify whether the benefit varies systematically with experience and whether repeated AI exposure produces a sustained training effect.

In conclusion, an integrated workflow combining automated DAB-TBA processing, digital whole-slide imaging, and interpretable deep learning showed high prospective discrimination for expert-consensus TBA positivity and higher reader performance in the AI-assisted round, with the largest observed difference among less-experienced readers. CNA-pattern subclassification remained comparatively challenging, and the current evidence does not establish independent multicentre or cross-platform generalizability. Further prospective validation is required before broader clinical implementation.

## Data availability

The de-identified data supporting the findings of this study are available from the corresponding authors upon reasonable request for bona fide research purposes, subject to approval by the relevant ethics committees and institutional data-governance requirements. Raw DAB-TBA images are not publicly available because they derive from clinical specimens and are subject to patient-privacy and institutional data-sharing restrictions. Access may require a data-use agreement. The custom code used for model training, inference, performance analysis, and Grad-CAM generation is available from the corresponding authors upon reasonable request, subject to institutional review and applicable use restrictions.

## Acknowledgements

During manuscript preparation, the authors used ChatGPT (OpenAI, GPT-5.6 Sol) to assist with English-language editing, sentence restructuring, manuscript organization, and formatting. All AI-assisted revisions were reviewed and approved by the authors, who take full responsibility for the final content.

## Author contributions

Z.C. and S.H.Z. conceived the study and designed the research. Z.C. and D.X. performed the AI model development and statistical analyses. Z.C., S.J.Z., and M.L. conducted the experiments, TBA testing, and data collection. D.L. and P.M.-M. contributed to clinical and professional interpretation. S.H.Z. coordinated and conducted the reader study. Z.C. drafted the original manuscript. D.X., S.H.Z., and Z.Z. critically reviewed and revised the manuscript. S.H.Z. and Z.Z. supervised the project and provided funding and resources. All authors reviewed and approved the final manuscript.

