## supplementary_material for "AI-Assisted Tissue-Based Antibody Interpretation in Autoimmune Encephalitis: Prospective Validation and Reader Study"

Zijian Cao et al.

### Supplementary figures

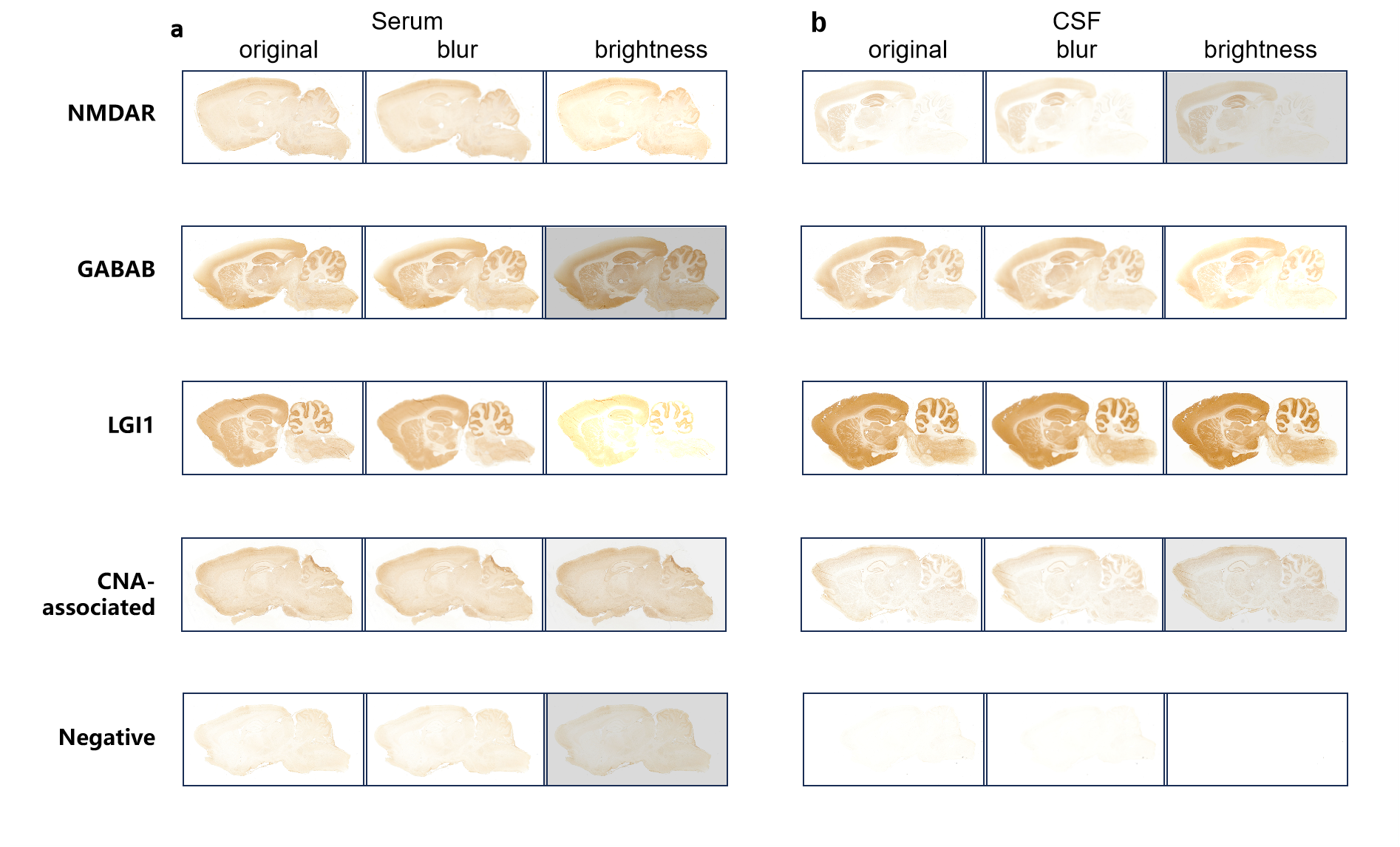

**Supplementary Figure 1.** Representative image variants used for augmentation of the internal DAB-TBA dataset. Representative serum and cerebrospinal fluid (CSF) images illustrate the fixed three-view representation: original image, mild Gaussian blur, and brightness/contrast adjustment. NMDAR-, GABABR-, LGI1-, CNA-associated, and negative examples are shown. Prospective validation used original images only. CNA, cytoplasmic/nuclear antigen-associated; DAB-TBA, 3,3′-diaminobenzidine tissue-based assay.

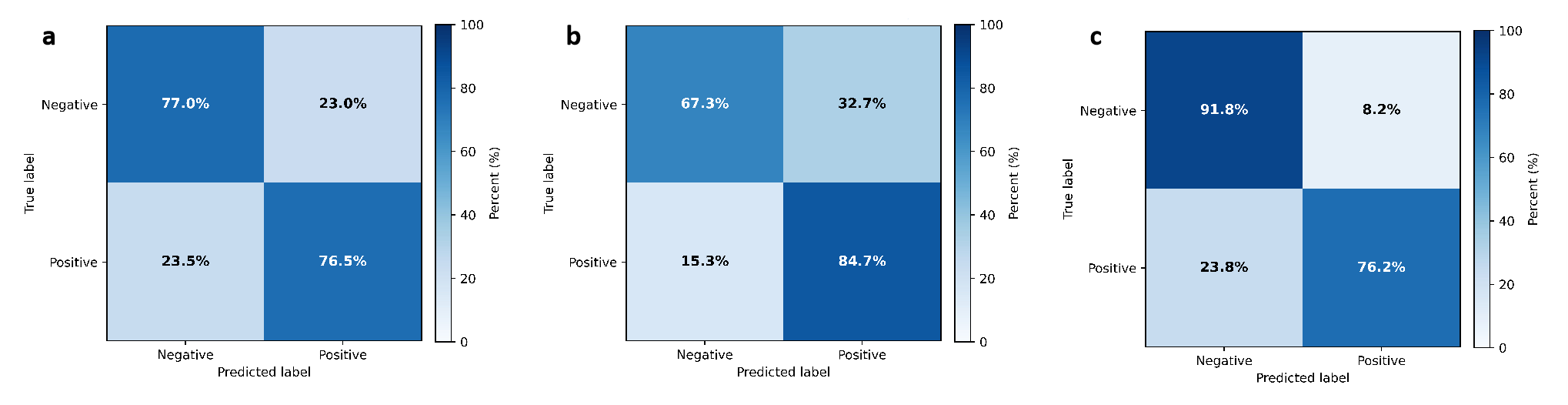

**Supplementary Figure 2.** Confusion matrices for internal binary TBA positive/negative classification. (A) Overall internal test set: sensitivity 76.5%, specificity 77.0%. (B) Serum: sensitivity 84.7%, specificity 67.3%. (C) Cerebrospinal fluid: sensitivity 76.2%, specificity 91.8%. Values are row-normalized percentages within reference-label categories.

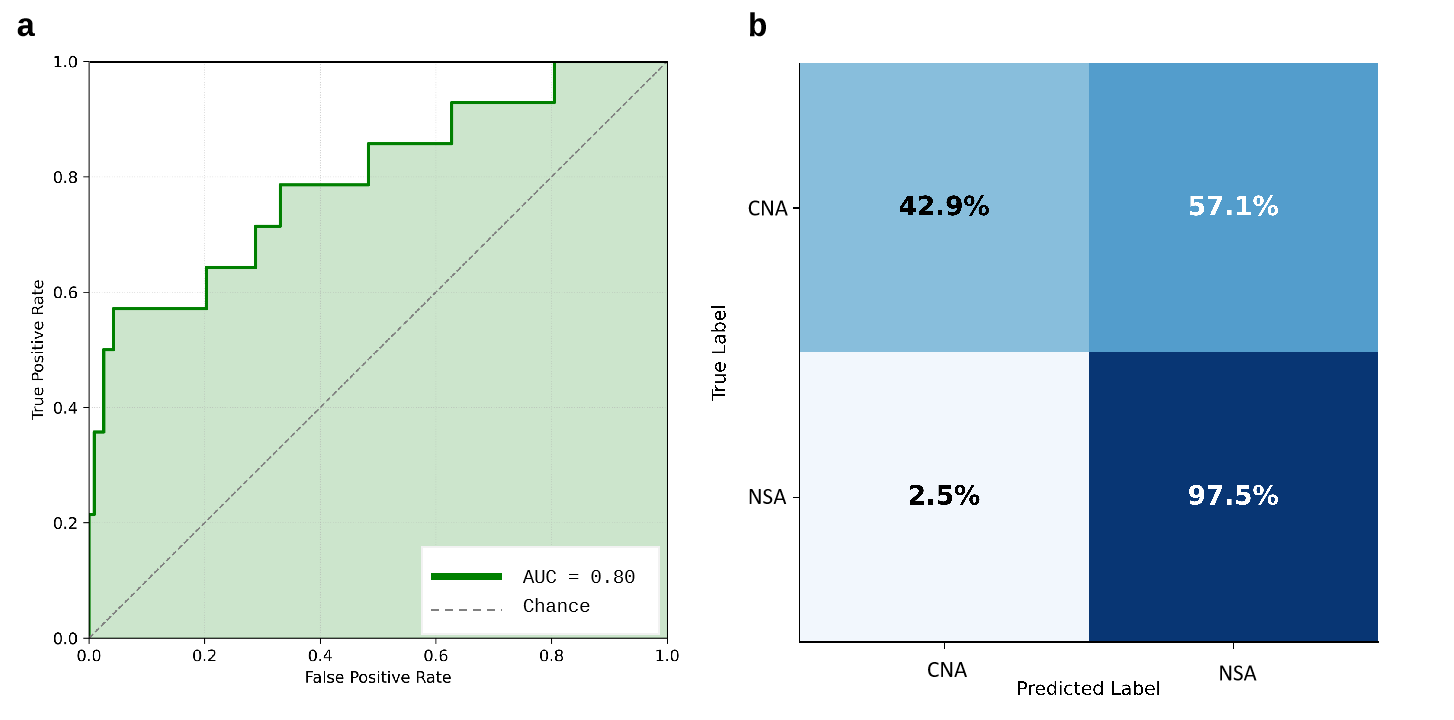

**Supplementary Figure 3.** Internal performance for CNA-versus-NSA staining-pattern subclassification among TBA-positive cases. The receiver operating characteristic curve shows AUC 0.797. The row-normalized confusion matrix shows CNA sensitivity 42.9% and NSA recall 97.5%. Analysis included 396 inference views derived from 132 TBA-positive original images. CNA, cytoplasmic/nuclear antigen-associated; NSA, neural surface antigen-associated.

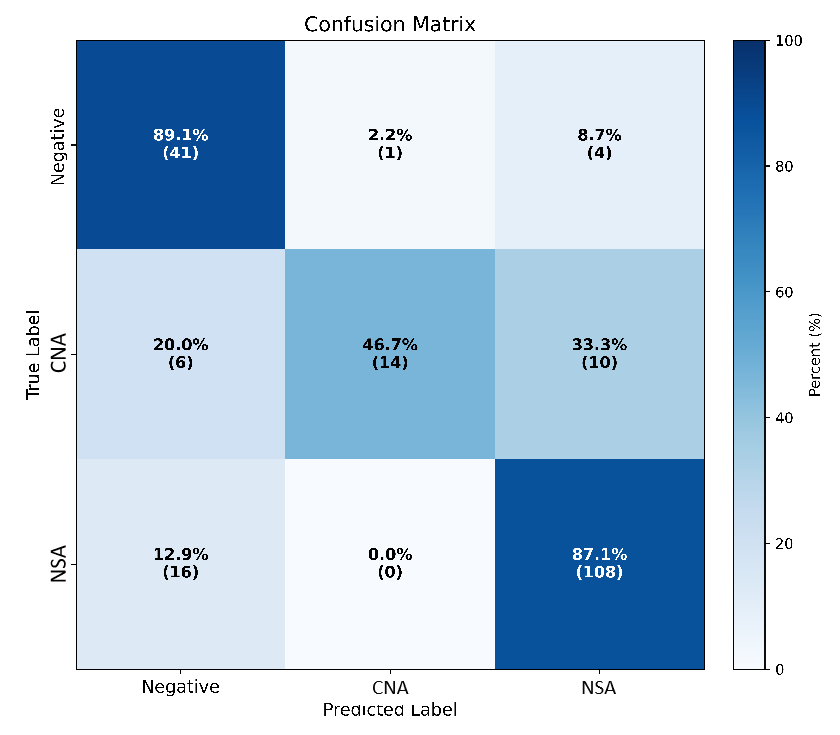

**Supplementary Figure 4.** Confusion matrix for end-to-end three-class classification in the prospective validation cohort. The three output classes were negative, CNA pattern, and NSA pattern. Overall end-to-end accuracy was 81.5% across 200 original images. Rows represent reference labels and columns represent model predictions. CNA, cytoplasmic/nuclear antigen-associated; NSA, neural surface antigen-associated.

### Supplementary tables

**Supplementary Table 1. End-to-end three-class cascade performance in the prospective validation cohort (n = 200)**

| **Class** | **Support** | **PPV** | **Recall** | **F1-score** |
| --- | --- | --- | --- | --- |
| Negative | 46 | 65.1% | 89.1% | 0.752 |
| CNA pattern | 30 | 93.3% | 46.7% | 0.622 |
| NSA pattern | 124 | 88.5% | 87.1% | 0.878 |
| Macro average | 200 | 82.3% | 74.3% | 0.751 |
| Weighted average | 200 | 83.9% | 81.5% | 0.811 |
| Overall accuracy | 200 | — | 81.5% | — |

Note. For end-to-end inference, images classified as positive by Stage 1 were passed to Stage 2, whereas images classified as negative by Stage 1 retained the negative label. The final cascade therefore produced one of three outputs: negative, CNA pattern, or NSA pattern. CNA, cytoplasmic/nuclear antigen-associated; NSA, neural surface antigen-associated.

**Supplementary Table 2. Conditional CNA-versus-NSA staining-pattern subclassification performance in the internal test set**

| **Subgroup** | **n (analysis units)** | **Accuracy** | **AUC** | **CNA sensitivity** | **NSA recall** | **Macro F1** |
| --- | --- | --- | --- | --- | --- | --- |
| Reference-standard TBA-positive cases | 396 inference views (132 originals) | 91.7% | 0.797 | 42.9% | 97.5% | 0.738 |

Note. This analysis was conditional on reference-standard TBA positivity and was calculated at the inference-view level. It should be distinguished from the end-to-end three-class cascade reported in Supplementary Table 1. CNA, cytoplasmic/nuclear antigen-associated; NSA, neural surface antigen-associated; TBA, tissue-based assay.

**Supplementary Table 3. AI-assisted reader study: binary TBA positive/negative interpretation by reading mode and specimen type**

| **Reading mode** | **Accuracy** | **Sensitivity** | **Specificity** | **Kappa (κ)** | **P value vs Round 1** |
| --- | --- | --- | --- | --- | --- |
| Overall cohort (n = 200) |  |  |  |  |  |
| AI only | 86.5% | 85.7% | 89.1% | 0.663 | — |
| Round 1 (unaided) | 90.4% | 92.7% | 82.6% | 0.735 | — |
| Round 2 (AI-assisted) | 97.4% | 96.8% | 99.5% | 0.928 | <0.001 |
| Serum subgroup (n = 100) |  |  |  |  |  |
| AI only | 92.0% | 96.1% | 79.2% | 0.774 | — |
| Round 1 (unaided) | 90.0% | 92.4% | 82.3% | 0.732 | — |
| Round 2 (AI-assisted) | 95.8% | 94.7% | 99.0% | 0.889 | <0.001 |
| CSF subgroup (n = 100) |  |  |  |  |  |
| AI only | 81.0% | 75.6% | 100.0% | 0.577 | — |
| Round 1 (unaided) | 90.8% | 92.9% | 83.0% | 0.738 | — |
| Round 2 (AI-assisted) | 99.0% | 98.7% | 100.0% | 0.971 | <0.001 |

Note. Round 1 and Round 2 values are pooled across four readers. The overall cohort therefore comprised 800 reader-image decisions per round, and each specimen subgroup comprised 400 reader-image decisions per round. P values were calculated using the paired McNemar test. Serum/CSF subgroup analyses were exploratory and P values are nominal two-sided values without additional multiplicity adjustment. Cohen’s κ denotes agreement with the reference-standard labels, not inter-reader agreement. CSF, cerebrospinal fluid; TBA, tissue-based assay.

**Supplementary Table 4. AI-assisted reader study: CNA-versus-NSA staining-pattern subclassification by reading mode and specimen type**

| **Reading mode** | **Accuracy** | **CNA sensitivity** | **NSA recall** | **Kappa (κ)** | **P value vs Round 1** |
| --- | --- | --- | --- | --- | --- |
| All TBA-positive cases (n = 154) |  |  |  |  |  |
| AI only | 79.2% | 46.7% | 87.1% | 0.431 | — |
| Round 1 (unaided) | 88.8% | 83.3% | 90.1% | 0.699 | — |
| Round 2 (AI-assisted) | 94.0% | 85.0% | 96.2% | 0.822 | <0.001 |
| Serum-positive subgroup (n = 76) |  |  |  |  |  |
| AI only | 85.5% | 37.5% | 98.3% | 0.496 | — |
| Round 1 (unaided) | 89.1% | 81.3% | 91.3% | 0.716 | — |
| Round 2 (AI-assisted) | 93.4% | 76.6% | 97.9% | 0.813 | 0.035 |
| CSF-positive subgroup (n = 78) |  |  |  |  |  |
| AI only | 73.1% | 57.1% | 76.6% | 0.395 | — |
| Round 1 (unaided) | 88.5% | 85.7% | 89.1% | 0.682 | — |
| Round 2 (AI-assisted) | 94.6% | 94.6% | 94.5% | 0.832 | <0.001 |

Note. Analysis was restricted to reference-standard TBA-positive cases. Round 1 and Round 2 values are pooled across four readers, corresponding to 616 reader-image decisions per round overall, 304 per round in the serum-positive subgroup, and 312 per round in the CSF-positive subgroup. CNA sensitivity denotes recall for the CNA class; NSA recall denotes recall for the NSA class. P values were calculated using the paired McNemar test. Specimen-type subgroup analyses were exploratory and P values are nominal. Cohen’s κ denotes agreement with the reference-standard labels. CNA, cytoplasmic/nuclear antigen-associated; NSA, neural surface antigen-associated; CSF, cerebrospinal fluid; TBA, tissue-based assay.

**Supplementary Table 5. Reader-experience analysis: binary TBA positive/negative interpretation**

| **Group** | **Reading mode** | **Accuracy** | **Sensitivity** | **Specificity** | **Kappa (κ)** | **P value vs Round 1** |
| --- | --- | --- | --- | --- | --- | --- |
| Reference | AI only | 86.5% | 85.7% | 89.1% | 0.663 | — |
| Expert | Round 1 (unaided) | 97.5% | 98.4% | 94.6% | 0.929 | — |
|  | Round 2 (AI-assisted) | 98.3% | 97.7% | 100.0% | 0.952 | 0.508 |
| Novice | Round 1 (unaided) | 83.3% | 87.0% | 70.7% | 0.549 | — |
|  | Round 2 (AI-assisted) | 96.5% | 95.8% | 98.9% | 0.906 | <0.001 |

Note. Expert readers had ≥5 years of dedicated TBA interpretation experience (n = 2); novice readers had <1 year of experience (n = 2). Expert and novice values are pooled across two readers (400 reader-image decisions per round in each experience group). P values are from paired McNemar tests comparing Round 2 with Round 1 within the same experience group. A Bonferroni-corrected significance threshold of α = 0.0125 was applied to the four reader-experience comparisons across Supplementary Tables 5 and 6. Cohen’s κ denotes agreement with the reference-standard labels. TBA, tissue-based assay.

**Supplementary Table 6. Reader-experience analysis: CNA-versus-NSA staining-pattern subclassification**

| **Group** | **Reading mode** | **Accuracy** | **CNA sensitivity** | **NSA recall** | **Kappa (κ)** | **P value vs Round 1** |
| --- | --- | --- | --- | --- | --- | --- |
| Reference | AI only | 79.2% | 46.7% | 87.1% | 0.431 | — |
| Expert | Round 1 (unaided) | 96.4% | 88.3% | 98.4% | 0.889 | — |
|  | Round 2 (AI-assisted) | 96.4% | 90.0% | 98.0% | 0.891 | 1.000 |
| Novice | Round 1 (unaided) | 81.2% | 78.3% | 81.9% | 0.556 | — |
|  | Round 2 (AI-assisted) | 91.6% | 80.0% | 94.4% | 0.756 | <0.001 |

Note. Analysis was restricted to the 154 reference-standard TBA-positive cases. Expert and novice values are pooled across two readers (308 reader-image decisions per round in each experience group). CNA sensitivity denotes recall for the CNA class; NSA recall denotes recall for the NSA class. P values are from paired McNemar tests comparing Round 2 with Round 1 within the same experience group. A Bonferroni-corrected significance threshold of α = 0.0125 was applied to the four reader-experience comparisons across Supplementary Tables 5 and 6. Cohen’s κ denotes agreement with the reference-standard labels. CNA, cytoplasmic/nuclear antigen-associated; NSA, neural surface antigen-associated; TBA, tissue-based assay.
